# SARS-CoV-2 post-vaccine surveillance studies in Australian children and adults with cancer: SerOzNET Quality of Life, Toxicity and Vaccine Beliefs Substudy Statistical Analysis Plan

**DOI:** 10.1101/2023.02.28.23286595

**Authors:** Mark W Donoghue, Amy Body, Claire Wakefield, Elizabeth Ahern, Eva Segelov

## Abstract

COVID-19 disease is associated with higher morbidity and mortality in cancer patients. SerOzNET is a prospective cohort study of adults and children with cancer undergoing routine SARS-CoV-2 vaccination in Australia. Peripheral blood was collected and processed at multiple points (one pre-vaccination and five or more post-vaccination) to address the primary aim of the study to assess the serological and immunological responses to vaccination.

A secondary aim of the study was to “document patient response to vaccination using qualitative measures, including patient-reported outcomes, vaccine hesitancy survey and post-hoc toxicity recording” (body, et al.,2022). This statistical analysis plan describes the analysis of the data collected to address this aim. We will refer to this as the SerOzNET QoL Substudy.

We have no conflicts of interest to disclose.

## 1 ADMINISTRATIVE INFORMATION

### 1.1 STUDY IDENTIFIERS

- Protocol: v6.0, 11 October 2021
  - (Body, et al., 2022)
- ANZCTR registration number: [ACTRN12621001004853](https://anzctr.org.au/Trial/Registration/TrialReview.aspx?id=382281&isReview=true)

Cancer Australia, Department of Health and Human Services Victoria, and Leukaemia Foundation have provided funding that partially supports this study. Cancer Australia contributed initial suggestions regarding project scope and focus. The authors subsequently independently developed and implemented the study protocol. Data analysis will be conducted independently without input from Cancer Australia. Department of Health and Human Services Victoria, and the Leukaemia Foundation have provided funding only with no role in study design, conduct or analysis.

### 1.2 REVISION HISTORY

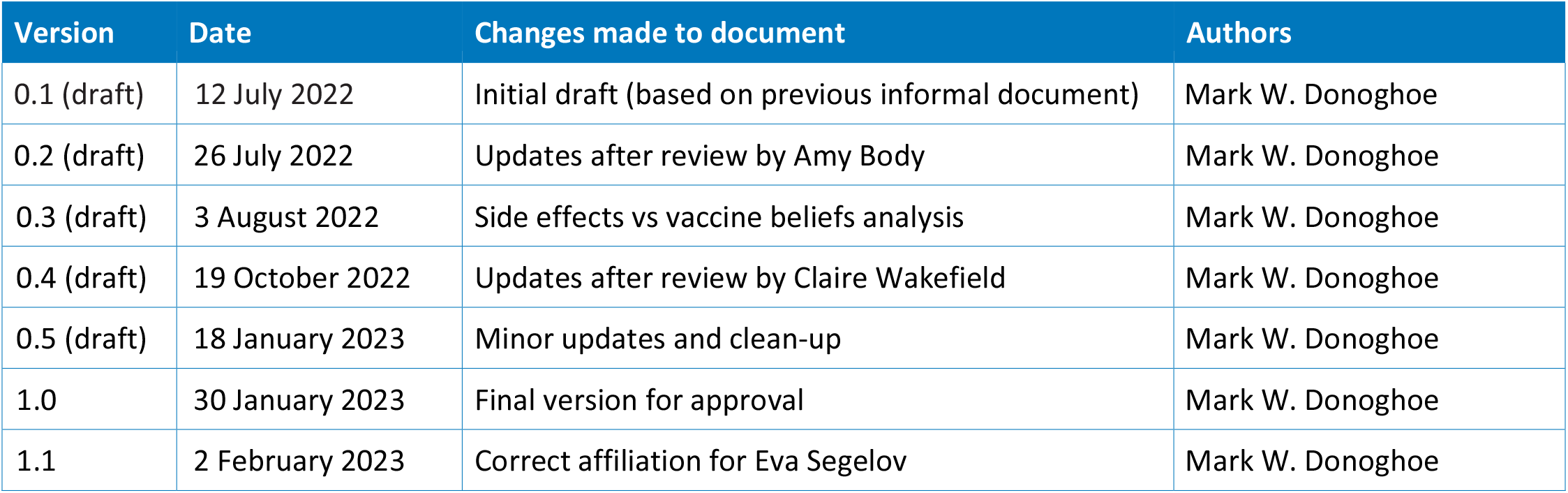

| Version | Date | Changes made to document | Authors |
| --- | --- | --- | --- |
| 0.1 (draft) | 12 July 2022 | Initial draft (based on previous informal document) | Mark W. Donoghoe |
| 0.2 (draft) | 26 July 2022 | Updates after review by Amy Body | Mark W. Donoghoe |
| 0.3 (draft) | 3 August 2022 | Side effects vs vaccine beliefs analysis | Mark W. Donoghoe |
| 0.4 (draft) | 19 October 2022 | Updates after review by Claire Wakefield | Mark W. Donoghoe |
| 0.5 (draft) | 18 January 2023 | Minor updates and clean-up | Mark W. Donoghoe |
| 1.0 | 30 January 2023 | Final version for approval | Mark W. Donoghoe |
| 1.1 | 2 February 2023 | Correct affiliation for Eva Segelov | Mark W. Donoghoe |

### 1.3 CONTRIBUTORS TO THE STATISTICAL ANALYSIS PLAN

#### 1.3.1 ROLES AND RESPONSIBILITIES

1.3 CONTRIBUTORS TO THE STATISTICAL ANALYSIS PLAN 1.3.1 ROLES AND RESPONSIBILITIES
| Name | Affiliation | Role on study | SAP contribution |
| --- | --- | --- | --- |
| Mark W. Donoghoe | Stats Central, Mark Wainwright Analytical Centre, UNSW Sydney | Statistician for this substudy | Main authorship |
| Amy Body | Monash Health and Monash University | Associate Investigator for SerOzNET; Principal Investigator for this substudy | Review and approval |
| Eva Segelov | Monash University and Faculty of Medicine University of Bern and University Cancer Centre, Inselspital | Principal Investigator for SerOzNET | Review and approval |
| Claire Wakefield | Behavioural Sciences Unit, Kids Cancer Centre, Sydney Children's Hospital and Discipline of Paediatrics and Child Health, School of Clinical Medicine, UNSW Sydney | Associate Investigator for this substudy | Review and approval |

#### 1.3.2 APPROVALS

The undersigned have reviewed this plan and approve it as final.

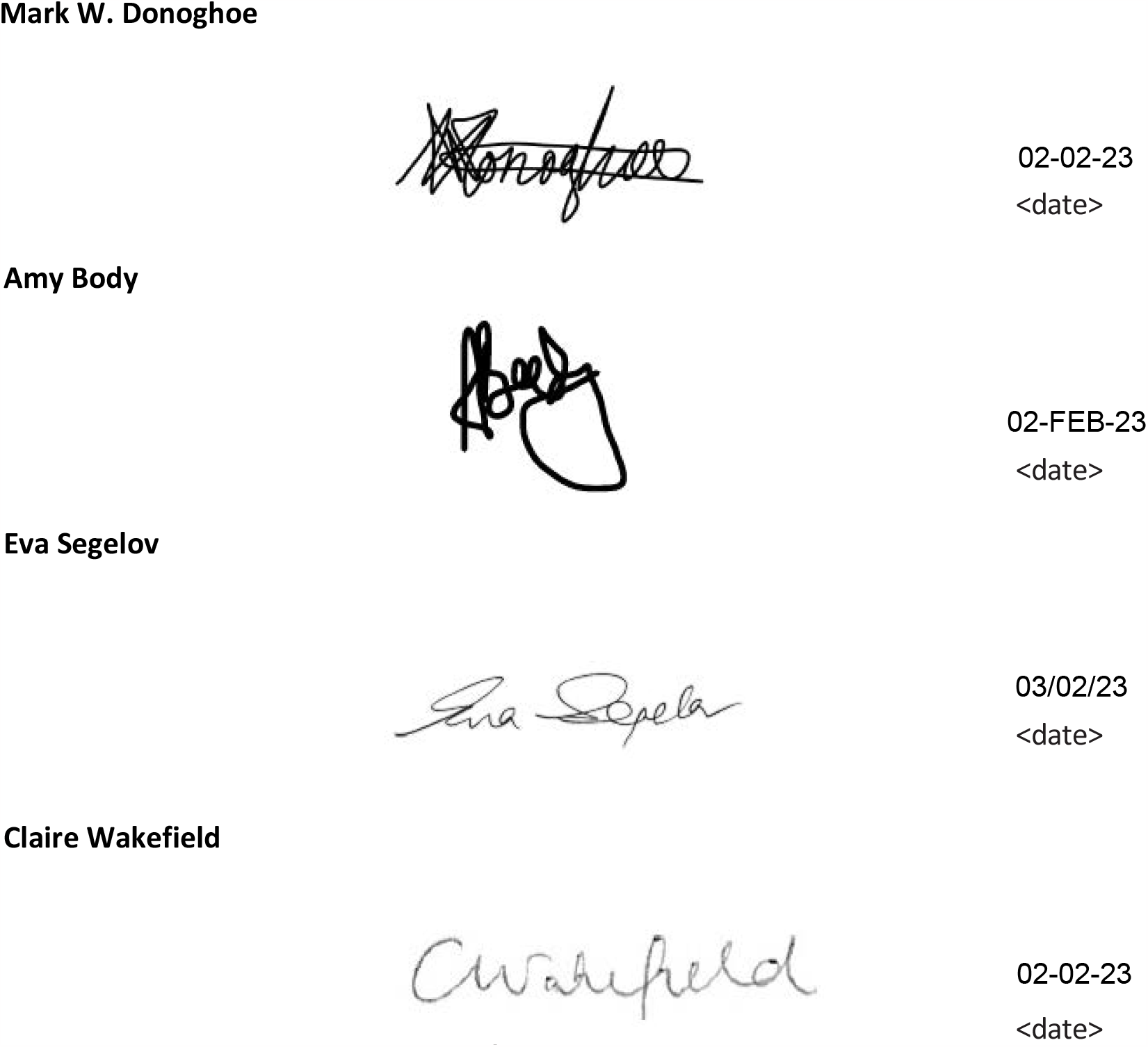

## 2 STUDY SYNOPSIS

SerOzNET is a prospective cohort study of adults and children with cancer undergoing routine SARS-CoV-2 vaccination in Australia. Peripheral blood was collected and processed at multiple points (one pre-vaccination and five or more post-vaccination) to address the primary aim of the study to assess the serological and immunological responses to vaccination.

A secondary aim of the study was to “document patient response to vaccination using qualitative measures, including patient-reported outcomes, vaccine hesitancy survey and post-hoc toxicity recording” (Body, et al., 2022). This statistical analysis plan describes the analysis of the data collected to address this aim. We will refer to this as the **QoL Substudy**.

### 2.1 QoL SUBSTUDY OBJECTIVES

#### 2.1.1 PRIMARY OBJECTIVE

Describe the safety and toxicity of COVID-19 vaccinations for patients with cancer. Specifically:

1. Describe detailed patient-reported and medically ascertained safety outcomes from COVID-19 vaccination for patients with cancer.
2. Assess whether side effects from vaccinations are associated with delays or interruptions to cancer treatment.
3. Assess whether patient-reported side effects from vaccinations are associated with increased vaccine-related concerns prior to the third dose.

#### 2.1.2 SECONDARY OBJECTIVES

1. Assess whether there are differences in the patient-reported and medically ascertained adverse event profile between vaccine types received by patients with cancer.
2. Describe the relationships between vaccine-related beliefs, quality of life, and toxicity. Specifically, assess whether self-reported adverse events and vaccine-related beliefs are associated with the following aspects of quality of life:

- Adults:
  - Emotional functioning
  - Global health status
  - Summary score
- Children and adolescents:
  - Worry
  - Procedural anxiety
  - Treatment anxiety
  - Communication
  - Total score

### 2.2 PATIENT POPULATION

Each of the analyses described in this plan will be performed separately within cohorts of patients defined by their age at the time of consenting to SerOzNET. Children will be defined as those aged between 5–11 (inclusive), adolescents will be defined as those aged between 12–19 (inclusive), and adults as those aged 20 and above.

#### 2.2.1 KEY INCLUSION CRITERIA

- Aged 5 years or over
- Able to give informed consent or legally provide consent on behalf of a person below 18 years of age
- Eligible for government COVID-19 vaccination program
- Cancer diagnosis fitting one of the study cohorts

#### 2.2.2 KEY EXCLUSION CRITERIA

- Not appropriate to have serial peripheral blood collections, for clinical reasons, e.g., severe anemia, poor venous access
- Life expectancy < 12 months

#### 2.2.3 PATIENT CHARACTERISTICS

The following patient characteristics were extracted from the medical record: date of birth, sex, Aboriginal or Torres Strait Islander status, country of birth, and languages spoken. Ethnicity was self-reported.

### 2.3 OUTCOMES

Patients (and/or their parent, guardian or caregiver, in the case of children with cancer) completed quality of life surveys prior to receiving their first and second vaccine doses, seven days after their first, second and third doses, and one month after their third dose (or three months after their second dose, if they didn’t have a third dose).

Patients (and/or their parent, guardian or caregiver, in the case of children with cancer) completed questions about their vaccine-related beliefs prior to receiving their first vaccine dose. Parents/guardians/caregivers also answered the same questions about their own beliefs, separate to the patient’s beliefs. Prior to receiving their third dose, adult patients completed these questions again, albeit with some of the questions modified for relevance.

Patients (and/or their parent, guardian or caregiver, in the case of children with cancer) reported adverse events via a PRO CTCAE survey seven days after their first, second and third doses, and study investigators completed a review of medical records to report any adverse events up until one month after a patient’s third vaccine dose.

#### 2.3.1 QUALITY OF LIFE OUTCOMES

##### EORTC QLQ-C30

Adult patients completed the EORTC QLQ-C30 questionnaire, version 3.0 (Aaronson, et al., 1993).

This comprises 28 questions rated on a four-point scale, which constitute five functional and nine symptom scales:

- Physical functioning (Questions 1–5)
- Role functioning (Q 6–7)
- Emotional functioning (Q 21–24)
- Cognitive functioning (Q 20, 25)
- Social functioning (Q 26–27)
- Fatigue (Q 10, 12, 18)
- Nausea and vomiting (Q 14–15)
- Pain (Q 9, 19)
- Dyspnoea (Q 8)
- Insomnia (Q 11)
- Appetite loss (Q 13)
- Constipation (Q 16)
- Diarrhoea (Q 17)
- Financial difficulties (Q 28)

There are an additional two questions, each rated on a seven-point scale, that comprise the “global health status / QoL” scale (Q 29–30). A *raw score* for each of these scales is calculated by averaging the responses to its corresponding questions, as long as at least half of those responses are non-missing. This is then transformed onto a 0–100 range, with functional scales reversed such that a high score:

- for a functional scale represents a high / healthy level of functioning,
- for the global health status / QoL scale represents a high QoL, but
- for a symptom scale represents a high level of symptomatology / problems.

Full details can be found in (Fayers, et al., 2001). Note that the majority of the survey asks the participant to respond with reference to the past week. The exceptions are questions 1–5 (which make up the physical functioning scale), which ask about current status: e.g., “Do you have trouble taking a long walk?” (Q 2).

The overall summary score is calculated by taking the mean of 13 of the 15 scales (the global health status / QoL and financial difficulties scales are not included), with symptom scales reversed. It is only calculated if all 13 scale scores are available (European Organisation for Research and Treatment of Cancer, 2018). A higher summary score represents better quality of life.

##### PedsQL

Children aged 8 and above and adolescents completed the PedsQL Cancer Module (Varni, Burwinkle, Katz, Meeske, & Dickinson, 2002), with the version depending on the child’s age:

- Age 8–12y: PedsQL Cancer Module v4.0 – Child report
- Age 13–18y: PedsQL Cancer Module v4.0 – Teen report

Parents of child and adolescent patients also completed proxy versions of the PedsQL Cancer Module, again depending on the child’s age:

- Age 5–7y: PedsQL Cancer Module v3.0 – Parent report for young children
- Age 8–12y: PedsQL Cancer Module v3.0 – Parent report for children
- Age 13–18y: PedsQL Cancer Module v3.0 – Parent report for teenagers

These questionnaires each follow the same format, comprising 26 or 27 questions (the parent report for young children excludes one question about essays and school projects). Each question is rated on a five-point scale, ranging from 0 (never) to 4 (almost always). These questions constitute eight dimensions:

- Pain and hurt (Questions 1–2)
- Nausea (Q 3–7)
- Procedural anxiety (Q 8–10)
- Treatment anxiety (Q 11–13)
- Worry (Q 14–16)
- Cognitive problems (Q 17–21 for ages 8+; Q 17–20 for ages 5–7)
- Perceived physical appearance (Q 22–24 for ages 8+; Q 21–23 for ages 5–7)
- Communication (Q 25–27 for ages 8+; Q 24–26 for ages 5–7)

A score for each question is calculated by reverse scoring the responses and linearly transforming onto a 0–100 range (such that 4 = 0; 3 = 25; 2 = 50; 3 = 75; 0 = 100). The scale score for each dimension is calculated by averaging the scores corresponding to that dimension, as long as at least half of those responses are non-missing. Higher scores indicate lower problems (Varni, Scaling and Scoring of the Pediatric Quality of Life Inventory).

A total score is calculated by averaging across all questions, noting that the questions—rather than the dimensions—are equally weighted. The scoring guide does not explicitly say that the total score is only calculated if at least half of the responses are non-missing, but we will apply this rule for consistency with the method of calculation used for the scale scores.

Note that this survey asks the participant to respond with reference to the past month, meaning that it could be less sensitive to short-term changes within the one-week window between pre- and post-vaccine time points.

#### 2.3.2 VACCINE-RELATED BELIEF OUTCOMES

The Oxford vaccine confidence and complacency scale (Freeman, et al., 2021) comprises 14 questions constituting four subscales that address vaccine-related beliefs about:

- Collective importance of a COVID-19 vaccine (Questions 4–5, 8, 11–12)
- Belief that the respondent may get COVID-19 and the vaccine will work (Q 1–3)
- Speed of vaccine development (Q 6–7, 13)
- Side effects (Q 9–10, 14)

Each question is rated on a five-point scale, with the additional option to respond “Don’t know”. The subscale score is calculated by averaging the responses corresponding to that subscale (excluding any “Don’t know” responses), as long as at least half of those responses are non-missing. For ease of interpretation, we will linearly rescale these scores onto a 0– 100 range. Higher scores indicate a greater degree of negative attitudes towards vaccination.

A total score can be calculated by averaging these subscale scores, as long as all of them are non-missing. This is not explicitly described in a previous publication, but was suggested in a personal communication by Aiden Lowe, who performed the statistical analysis for the scale development.

The three questions related to the speed of vaccine development were not included in the questionnaire at dose 3, and were replaced by three alternative questions that were more relevant to the booster dose. These alternative questions had different response formats to the remaining questions and hence could not be summarised by a subscale score in the same way. Given this, in order to compare total scores between dose 1 and dose 3, we calculate a total score that excludes the speed subscale. The additional three questions will be used for descriptive purposes only.

#### 2.3.3 PATIENT-REPORTED SIDE EFFECTS AND ADVERSE EVENTS

The PRO CTCAE questionnaire included questions that were specifically relevant to vaccine-related adverse events. The questions had several different formats and response options. In the case of children with cancer, the patient’s parent, guardian or caregiver could answer this questionnaire on behalf of the child; throughout this document we will still refer to these as “patient-reported” outcomes.

The listed side effects and adverse events were grouped into five overarching categories:

- Local side effects:
  - Pain at the vaccine injection site‡
  - Redness or swelling at the injection site†
  - Itch at the injection site†
- Systemic side effects:
  - Fever after the vaccination†
  - New or worsened fatigue‡
  - New or worsened headache†
  - Shivering or shaking chills*
  - New or worsened muscle pain†
  - New or worsened joint pain‡
  - New or worsened nausea†*
  - New or worsened vomiting†
  - New or worsened diarrhoea†*
- Other side effects:
  - Medications taken to treat side effects
  - New treatment for blood clot
  - Any other side effects (free text)
- Impact on cancer treatment:
  - Having to delay or stop cancer treatment
- Healthcare utilisation:
  - Emergency department visits
  - Hospital admissions
  - Local doctor visits

Questions marked with † also asked the patient to describe the severity of the side effect, those marked with ‡ asked the patient to describe how much it interfered with their usual or daily activities, and those marked with * asked the patient to describe the frequency of the side effect. Local and systemic side effect questions were taken directly from the published PRO-CTCAE library (National Cancer Institute). Other side effects, impact on cancer treatment and healthcare utilisation questions were developed by the investigators.

For each category and overall, at each time point we will calculate a binary variable to indicate if the patient reported an adverse event. We will also calculate a binary variable across all three categories relating to vaccine side effects. These binary variables will be “Yes” if the patient indicated that any of the given events occurred (even if some other responses are missing), they will be “No” if the patient indicated that none of the given events occurred (with no missing responses), and will be missing otherwise (i.e., if there are some missing responses and no “Yes” responses).

For side effects where either severity or interference with daily activities was collected (that is, all local and systemic side effects, except shivering or shaking chills), at each time point we will calculate an ordinal variable to describe the worst severity that the patient reported across these side effects. For the purposes of this summary, the five possible options for each side effect will be treated as equivalent, regardless of its label. For example, pain has options of [1] “No”, [2] “Mild”, [3] “Moderate”, [4] “Severe” and [5] “Serious”, and redness or swelling has options of [1] “No”, [2] “Minor”, [3] “Moderate”, [4] “Severe” and [5] “Serious”. A response of [2] on either question will be treated as the second-highest level of severity.

The “any other side effects” category will be considered as a “No” if the patient did not provide any free-text response. Otherwise the responses will be manually classified as additional local side effects, additional systemic side effects or invalid responses (e.g., free text not referring to a side effect, or repetition of a side effect that is already captured in the questionnaire). For local doctor visits, if the patient indicates that they visited only for a “routine check or prescription”, this will be treated as a “No” response for the purposes of calculating the summary binary variables.

#### 2.3.4 MEDICALLY ASCERTAINED ADVERSE EVENTS

A review of patients’ medical records was conducted, and collated data recorded by the health care team about any severe adverse events (SAEs) that occurred between baseline and one month after dose 2, as well as between dose 3 and one month after dose 3. The event type and grade (as per CTCAE) were recorded, along with an assessment of whether the event was attributable to the vaccine (yes / possibly / unlikely), according to the patient’s treating physician. If such an attribution was not recorded, it was decided by a study investigator.

Any cancer treatment delays and modifications in these time windows that were documented in medical records were also recorded, along with the reasons.

Thrombotic events, declines in performance status and lymphadenopathy in these time windows were also recorded.

Allergic reactions, and their types, were recorded for each vaccine dose.

### 2.4 VACCINATION PRODUCTS

Three different vaccine products were available during the study: AstraZeneca, Pfizer and Moderna. Comparisons between vaccine types will compare the viral vector AstraZeneca vaccine to the mRNA-based Pfizer and Moderna vaccines (combined into a single group). Because official advice recommended that people under 60 years old should receive an mRNA vaccine, these comparisons will only be performed in those aged 60 years and above. These comparisons will be performed separately at each time point, such that if a patient received different vaccines at different time points, they will be included in the appropriate group at each time point.

### 2.5 SAMPLE SIZE

The primary outcome of the main SerOzNET study is the seroconversion rate. According to the main study’s sample size calculation (Body, et al., 2022), within a subgroup defined by cancer type, 100 individuals would provide 80% power to detect a decrease of 10% seroconversion rate compared to an assumed rate of 95% in a non-cancer population. This suggests a total sample size of 600 patients.

The sample size for this substudy will be determined by the data available from the main study, and we have not performed a sample size or power calculation. All estimates will be presented with 95% confidence intervals in order to reflect the uncertainty that results from the available sample size.

## 3 STATISTICAL ANALYSIS

### 3.1 GENERAL PRINCIPLES

The scope of this analysis plan is to address the research questions related to quality of life, toxicity and vaccine-related beliefs, described in Section 2.1. Analysis of other aspects of the SerOzNET study are described separately.

The analysis plan for this substudy was previously outlined in an informal document. This document supersedes it, providing more rigorous descriptions of the planned analyses.

Analyses will be conducted using R (R Core Team, 2021), with the targets package (Landau, 2021) used to manage dependencies, git (Chacon & Straub, 2014) to implement version control, and knitr (Xie, Dynamic Documents with R and knitr, 2015) and R Markdown (Xie, Allaire, & Grolemund, R Markdown: The Definitive Guide, 2018) to dynamically generate reports.

Where relevant, parameter estimates will be presented with two-sided 95% confidence intervals. Two-sided p-values will be calculated for hypothesis tests. No formal adjustment for multiple comparisons will be performed, except where noted in this plan.

### 3.2 ANALYSIS POPULATION

The analysis population includes all individuals who were enrolled into SerOzNET and completed at least one adverse event or quality of life questionnaire, or had a medical record review. The population will be separated into three distinct cohorts based on the patient’s age at the time of consenting to SerOzNET: children (5–11 years, inclusive), adolescents (12–19 years, inclusive), and adults (20 years and above).

### 3.3 SUBJECT DISPOSITION

Individuals will be classified according to how many doses of the vaccine they received, and the questionnaires that they completed at each time point. A flowchart describing subject disposition in more detail will be created if necessary.

For each patient, we have a record of the dates on which they received each vaccination dose. For assessing QoL completion rates, we allow a ‘grace period’ after the scheduled date of 7 days for the post-dose 1 survey, and 14 days for the other surveys before considering a survey to be not completed. Given that the third dose of the vaccine was optional, where none has been recorded, we separate patients who have not had a third dose from those who may not be due to have their third dose based on whether it has been at least 4 months since their second dose (or 4 months + 8 weeks since their first dose, for those without a second dose date).

### 3.4 PATIENT CHARACTERISTICS

Patient characteristics will be summarised separately within each cohort and overall. Age at the time of consent will be calculated from the patient’s date of birth, and summarised by its mean and standard deviation, median and interquartile range, and minimum and maximum. The remaining collected characteristics—sex, country of birth (Australia/Other), Aboriginal or Torres Strait Islander status, and primary language spoken at home (English/Other)—will be summarised using frequencies and proportions.

### 3.5 ANALYSES ADDRESSING THE SUBSTUDY PRIMARY OBJECTIVE

Each of the questions that constitute the primary objective will be addressed with separate analyses, described in the sections below.

#### 3.5.1 PATIENT-REPORTED AND MEDICALLY ASCERTAINED SAFETY OUTCOMES

Patient-reported side effects and adverse events will be summarised by their incidence (number and proportion of patients) at each dose. This will be done individually for each listed side effect or adverse event, grouping over the categories described in Section 2.3.3, and overall.

Medically ascertained adverse events (SAEs, thrombotic events, declines in performance status, lymphadenopathy) and their recorded properties (e.g., grade, type and attribution to vaccine, for SAEs) will be summarised by their incidence in each time window (baseline to one month after dose 2, and dose 3 to one month after dose 3). Allergic reactions and their types will be summarised by their incidence at each dose.

#### 3.5.2 DELAYS AND INTERRUPTIONS TO CANCER TREATMENT

The incidence of delays and modifications to cancer treatment at each time point reported on medical record reviews will be summarised as the number and proportion of patients for whom these were recorded. To address this objective in particular, we will summarise the incidence of delays and modifications that were described as vaccine-related, along with the free-text reasons that were recorded. In order to avoid double-counting adverse events that resulted in both a delay and a modification, we will also summarise the number and proportion of individuals for whom either a delay *or* a modification (or both) for vaccine-related reasons was recorded at each time point.

To address this research question, we will create a binary variable grouping over the three categories of patient-reported side effects (local / systemic / other) to summarise the number and proportion of patients reporting any side effects after each dose. We will use Fisher’s exact test to examine if there is evidence of an association between this binary variable and patient-reported delays or interruptions to cancer treatment, separately at each dose.

The number of patients with missing data for each of these outcomes will be reported, but these patients will be excluded from the analyses.

#### 3.5.3 ASSOCIATION BETWEEN PATIENT-REPORTED SIDE EFFECTS AND VACCINE-RELATED BELIEFS

Each subscale and the total score of the Oxford vaccine confidence and complacency scale will be summarised at each time point using histograms, means and standard deviations and medians and interquartile ranges.

Patient-reported side effects prior to dose 3 will be based on the ordinal variables that describe the highest severity that the patient reported (as described in Section 2.3.3) after doses 1 and 2, summarised into a single ordinal variable describing the highest severity reported across both time points. If any level of severity has fewer than 10 observations, it will be combined with the adjacent level of severity that has the smaller number of observations.

The outcome of interest is the patient’s total vaccine-related beliefs score collected before dose 3. ANCOVA will be used to assess the evidence for an association between highest severity of side effects at the first two doses and this outcome, adjusting for the baseline vaccine-related beliefs score which excludes the speed subscale (see Section 2.3.2). Specifically, we will use an F-test to calculate a p-value for the interaction between the baseline score and side effects. If there is evidence of an interaction (p < 0.05), the results will be summarised by estimating the difference in outcome scores between patients with side effects of each level of severity, at varying values for the baseline score. If there is not strong evidence of an interaction (p > 0.05), we will repeat the analysis without an interaction term, to calculate a p-value for the ‘main effect’ of side effect severity. In this case, the estimated difference in outcome scores between patients with different severity of side effects does not depend on the baseline score.

If there is evidence of an association between side effect severity and the total vaccine-related beliefs score, the analyses will be repeated for the individual subscales (excluding speed).

We will summarise the association between both baseline vaccine-related beliefs (total score) and patient-reported side effects after doses 1 and 2 (highest severity, as described above) and whether a patient received dose 3 via separate logistic regression models. We anticipate that patients with more negative vaccine-related beliefs at baseline and those who experience more severe side effects after their first two doses may have been less likely to receive dose 3, meaning that the data will not be missing completely at random (MCAR), and a complete-case analysis will produce biased results.

Therefore, we will use predictive mean matching to create 25 imputed complete datasets (van Burren & Groothuis-Oudshoorn, 2011), based on patient age, gender, vaccine-related beliefs and patient-reported side effects. The analysis will be performed separately on each imputed dataset and the results combined (Rubin, 2004) (Grund, Lüdtke, & Robitzsch, 2016) to produce the overall result, which will be valid if the data are missing at random (MAR).

### 3.6 ANALYSES ADDRESSING SUBSTUDY SECONDARY OBJECTIVES

Each of the secondary objectives will be addressed with separate analyses, described in the sections below.

#### 3.6.1 ADVERSE EVENTS BY VACCINE TYPE

As described in Section 2.4, comparisons between vaccine types will be undertaken in patients over 60 years at consent to SerOzNET, and will group together the mRNA-based Pfizer and Moderna vaccines to compare against the viral vector AstraZeneca vaccine. Comparisons will be performed separately at each dose, with patients classified according to the vaccine type they received at the given time. Patients who did not receive a particular dose will not be included.

The frequency and proportion of patients reporting each type of side effect or adverse event at each dose will be summarised, and compared between vaccine types using Fisher’s exact tests. Where appropriate, the reported severity, impact and frequency of these side effects or adverse events will be analysed in the same way.

The incidence of adverse events reported on the medical record review, will be analysed in the same way, separately for doses 1–2 and dose 3. If there were any patients who received different vaccine types for their first and second doses, they will be excluded. Given that the follow-up time differed systematically between vaccine types (since the time between dose 1 and one month post-dose 2 depends on the gap between doses 1 and 2), these outcomes will also be analysed using a log-link Poisson generalised linear model (GLM), with an offset of log(follow-up time), in order to examine if there is evidence that the *rate* of adverse events differed between vaccine types. We will use a likelihood ratio test to calculate a p-value for a test of the difference in rates.

The number of serious adverse events reported for each patient at each time point will also be calculated, and analysed using log-link Poisson GLMs both with and without an offset for follow-up time, in order to examine differences in the expected number and rate of SAEs.

Estimates of the expected number and rate of events in each group from these models will be presented with 95% confidence intervals. If there is evidence of overdispersion in any of these models, a negative binomial GLM will be used instead.

#### 3.6.2 ASSOCIATION BETWEEN BASELINE QUALITY OF LIFE AND VACCINE-RELATED BELIEFS

For the second of the secondary objectives, the following QoL scales were identified as being of primary importance:

- Adults:
  - Emotional functioning
  - Global health status
  - Summary score
- Children and adolescents:
  - Worry
  - Procedural anxiety
  - Treatment anxiety
  - Communication
  - Total score

Only these will be examined in the analyses.

We will estimate Spearman’s correlation coefficient (and its 95% confidence interval) between the patient’s baseline total score of the Oxford vaccine confidence and complacency scale and the listed baseline QoL scales.

Scatterplots will be produced to visualise these associations. Patients with missing data for the vaccine beliefs score will be excluded from all analyses; those with missing data for QoL scales will be excluded from individual analyses as appropriate.

#### 3.6.3 ASSOCIATION BETWEEN QUALITY OF LIFE AND PATIENT-REPORTED SIDE EFFECTS OR ADVERSE EVENTS

We will fit separate logit-link binomial GAMs (i.e., logistic regression models) to estimate the relationship between the listed baseline QoL scores and the binary variable that summarises the incidence of *any* patient-reported side effects or adverse events after dose 1 or 2. We will plot the estimated relationship between the QoL scores and the probability of a side effect or adverse event, along with a 95% confidence interval. We will use a Wald test to calculate an approximate p-value for a test that the association is not zero (Wood, 2013).

Patients with missing data will be excluded from these analyses as appropriate, and the results will be valid under an assumption that data is missing completely at random. If the amount of missing data is substantial (> 5% of patients are excluded for a given analysis), we will conduct a sensitivity analysis using multiple imputation to assess the robustness of the results to the MCAR assumption.

## Data Availability

All data produced in the present study are available upon reasonable request to the authors.

## LIST OF ABBREVIATIONS

AE: Adverse event
COVID-19: Coronavirus disease 2019
EORTC: QLQ-C30 European Organisation for the Research and Treatment of Cancer Quality of Life Questionnaire
GAM: Generalised additive model
GLM: Generalised linear model
MAR: Missing at random
MCAR: Missing completely at random
mRNA: Messenger ribonucleic acid
PedsQL: Pediatric Quality of Life Inventory
PRO CTCAE: Patient-Reported Outcomes version of the Common Terminology Criteria for Adverse Events
QoL: Quality of life
SAE: Serious adverse event
SARS-CoV-2: Severe acute respiratory syndrome coronavirus 2

## REFERENCES

Aaronson, N. K., Ahmedzai, S., Bergman, B., Bullinger, M., Cull, A., Duez, N. J., … Takeda, F. (1993). The European Organisation for Research and Treatmetn of Cancer QLQ-C30: A quality-of-life instrument for use in international clinical trials in oncology. Journal of the National Cancer Institute, 85, 365–376.

Body, A., Ahern, E., Lal, L., Gillett, K., Abdulla, H., Opat, S., … Segelov, E. (2022). Protocol for SARS-CoV-2 post-vaccine surveillance study in Australian adults and children with cancer: an observational study of safety and serological and immunological response to SARS-CoV-2 vaccination (SerOzNET). BMC Infectious Diseases, 22, 70. doi:10.1186/s12879-021-07019-1

Chacon, S., & Straub, B. (2014). Pro git. Apress.

European Organisation for Research and Treatment of Cancer. (2018). Scoring of the QLQ-C30 Summary Score. Retrieved from https://qol.eortc.org/app/uploads/sites/2/2018/02/scoring_of_the_qlq-c30_summary_score.pdf

Fayers, P. M., Aaronson, N. K., Bjordal, K., Groenvold, M., Curran, D., & Bottomley, A. (2001). The EORTC QLQ-C30 Scoring Manual (3rd Edition). Brussels: European Organisation for Research and Treatment of Cancer.

Freeman, D., Loe, B. S., Chadwick, A., Vaccari, C., Waite, F., Rosebrock, L., … Lambe, S. (2021). COVID-19 vaccine hesitancy in the UK: the Oxford coronavirus explanations, attitudes, and narratives survey (Oceans) II. Psychological Medicine, First view, 1–15. doi:10.1017/S0033291720005188

Grund, S., Lüdtke, O., & Robitzsch, A. (2016). Pooling ANOVA results from multiply imputed datasets. Methodology, 12(3), 75–88. doi:10.1027/1614-2241/a000111

Landau, W. M. (2021). The targets R package: A dynamic Make-like function-oriented pipeline toolkit for reproducibility and high-performance computing. Journal of Open Source Software, 6(57), 2959. doi:10.21105/joss.02959

National Cancer Institute. (n.d.). PRO-CTCAE Measurement System. Retrieved from https://healthcaredelivery.cancer.gov/pro-ctcae/instrument-pro.html

R Core Team. (2021). R: A language and environment for statistical computing. Vienna: R Foundation for Statistical Computing. Retrieved from https://www.R-project.org/

Rubin, D. B. (2004). Multiple Imputation for Nonresponse in Surveys. John Wiley & Sons.

van Burren, S., & Groothuis-Oudshoorn, K. (2011). mice: Multivariate Imputation by Chained Equations in R. Journal of Statistical Software, 45(3), 1–67. doi:10.18637/jss.v045.i03

Varni, J. W. (n.d.). Scaling and Scoring of the Pediatric Quality of Life Inventory. Lyon: Mapi Research Trust. Retrieved from https://www.pedsql.org/PedsQL-Scoring.pdf

Varni, J. W., Burwinkle, T. M., Katz, E. R., Meeske, K., & Dickinson, P. (2002). The PedsQL in pediatric cancer: Reliability and validity of the pediatric quality of life inventory generic core scales, multidimensional fatigue scale, and cancer module. Cancer, 94(7), 2090–2106. doi:10.1002/cncr.10428

Wood, S. N. (2013). On p-values for smooth components of an extended generalized additive model. Biometrika, 100(1), 221–228. doi:10.1093/biomet/ass048

Xie, Y. (2015). Dynamic Documents with R and knitr (2nd edition ed.). Chapman and Hall/CRC.

Xie, Y., Allaire, J. J., & Grolemund, G. (2018). R Markdown: The Definitive Guide. Chapman and Hall/CRC. Retrieved from https://bookdown.org/yihui/rmarkdown

